# Susceptibility to monkeypox virus infection: seroprevalence of orthopoxvirus in 4 population samples; France, Bolivia, Laos and Mali

**DOI:** 10.1101/2022.07.15.22277661

**Authors:** Léa Luciani, Nathanaël Lapidus, Abdennour Amroun, Alessandra Falchi, Chanthala Souksakhone, Mayfong Mayxay, Audrey Dubot-Pérès, Paola Mariela Saba Villarroel, Issa Diarra, Ousmane Koita, Pierre Gallian, Xavier de Lamballerie

## Abstract

Europe is experiencing an epidemic of monkeypox virus against a background of limited knowledge of population immunity to orthopoxviruses. We tested antibodies neutralizing vaccinia virus in blood samples from Bolivia (n=601), Laos (n=657), Mali (n=255) and France (n=4876). We also tested neutralization of cowpox virus in 4448 French samples, which confirmed extensive cross-immunity between both viruses. Using a cut-off titer of 20, seroprevalence is <1% in Bolivia, <5% in Laos (mainly related to vaccination in the elderly), 17.25% in Mali (related to both smallpox vaccination and the likely natural circulation of orthopoxviruses). In France, neutralizing antibodies are found (but at low prevalence) in age groups supposed to be vaccinated, suggesting both decrease in antibody titer in vaccinated individuals and decline in smallpox vaccine compliance before the end of compulsory vaccination. We conclude that the populations tested in Europe, Africa, Asia and South-America are massively susceptible to orthopoxvirus (*e.g*. monkeypox) infections.

**Article summary line:** **S**eroprevalence of orthopoxvirus tested on 4 continents is very low, even for people born before the eradication of smallpox, making populations vulnerable to the emergence of zoonotic orthopoxviruses such as monkeypox virus.

## Introduction

In the context of the emergence of monkeypox virus (MPXV) infections, it is legitimate to question the immunity of populations to viruses of the genus *Orthopoxvirus*, to which MPXV, smallpox virus, and vaccinia virus belong. There is a broad cross-immunity between the viruses of this genus, which allowed the use of vaccinia virus as a vaccine to prevent smallpox and which today leads to the use of a vaccinia-derived vaccine to prevent or mitigate MPXV infections.

Since 1980, with the cessation of smallpox vaccination, the immunity of the world population against orthopoxviruses has decreased and this has been associated in young populations with the emergence of zoonotic orthopoxviruses with extended host specificity. This is the case of MPXV, which has been responsible for epidemic episodes in Africa *(1–3)* (which is its original territory of widespread) but also for cases outside this continent*(4,5)*. This is also the case for buffalopox virus*(6–8)* and camelpox virus*(8–10)* in Asia and for Cowpox virus, which is ubiquitous*(11–13)*. There is a clear risk that these orthopoxvirus emergence events will become more and more common due to increased travel and trade, ecosystem change, and altered biodiversity and climate*(14–16)*. Since the eradication of smallpox in 1980, work on orthopoxviruses in human medicine has gradually declined. However, in the early 2000’s much attention has been paid to the bioterrorism risk associated with smallpox*(17–19)*. This has led to attempts to assess the susceptibility of the general population to smallpox attacks*(18)*, which have generally been based on estimates of smallpox vaccination coverage. In France, Santé Publique France (the French Institute of Public Health) published a report in 2001 based on data from INSEE (National Institute of Statistics and Economic Studies) and Inserm (National Institute of Health and Medical Research), which estimated vaccination coverage in the French population close to 0% for persons born after 1979, 50% for those born between 1972 and 1978, 65% for those born between 1966 and 1971 and 90% for those born prior to 1966*(20)*. As in most developed countries, the strategy to fight smallpox in France has been based on systematic and mandatory vaccination of children. Vaccination consisted in two injections, the first at the age of one and the second usually around the age of 11. It was mandatory from 1902 to 1978 (until 1984 for the booster). However, for many resource-limited countries, routine vaccination of the population was difficult to implement, and the WHO shifted in the 1960s to a containment strategy of case identification, isolation, and widespread vaccination of contacts. As is well known, this strategy has been successful in eradicating smallpox*(21)*, but the vaccination coverage of the general population in these countries (which confers potential cross-immunity to other orthopoxviruses) is therefore likely to be lower than in countries where routine vaccination has been organized.

In this article, we present a large-scale epidemiological study based on the detection of antibodies neutralizing the vaccinia virus and the cowpox virus. We tested ∼6500 persons from four countries of different continents: France France and three low-income countries: Laos, Mali and Bolivia. In the current context of MPXV dissemination, these data provide a recent demographic overview of orthopoxviruses seroprevalence and allow assessment of the susceptibility of relevant populations to infection by this group of viruses.

## Materials and methods

### Study population and ethical approval

We investigated four different populations from France, Bolivia, Laos and Mali. All of them were tested for the presence of antibodies neutralizing the vaccinia virus. In addition, a large part of the French population was tested for the presence of antibodies neutralizing the cowpox virus.

i. The French population included 4876 voluntary - unpaid - blood donors, whose serum samples were collected in 2012, 2013, and 2019 from four regions of metropolitan France: Auvergne-Loire (n=837), Corsica (n=596), Midi-Pyrénées (n=1738), and Provence-Alpes-Côte d’Azur (PACA) (n=1705). Donors provided signed informed consent for the use of their blood samples for non-therapeutic research purposes. They completed a questionnaire that included year of birth, gender, and detailed information about their lifestyle, environment (home and workplace), and exposure to zoonotic diseases*(22)*.
ii. The Bolivian population included 601 voluntary, unpaid, blood donors*(23)*, whose serum samples were collected in 2017 among the main cities (urban environment) of 5 departments: Santa Cruz de la Sierra (n=165) and Beni (n=102) (tropical climate); Cochabamba (n=151), Tarija (n=23) and La Paz (n=160) (colder subtropical climate in the highlands). This study was approved by the ethics committee of the Medical College of Santa Cruz and donors provided signed informed consent for research use of their blood samples. The information collected included the year of birth, sex, city of residence and occupation.
iii. For Laos, collection of blood samples from 657 blood donors took place in the capital city of Vientiane in 2003, 2004, 2015 and 2018. Donors provided signed informed consent for research use of their blood samples and the study was approved by the Lao National Health Research Ethics Committee and the Oxford Tropical Research Ethics Committee. The information collected was limited to year of birth and gender.
iv. In Mali, 257 blood samples were collected in 2019 in the villages of Leba, Tliemba, Soloba, Bougoudale and Komana for a baseline study of health indicators in the villages of the Komana gold mine (tropical forest area). Participants provided informed consent for research use of their blood samples and the study was approved by the ethics committee of the National Institute for Public Health Research in Mali. The information collected was limited to year of birth and gender.

### Seroneutralization techniques

We used the Western Reserve vaccinia strain and the Compiègne strain of cowpox virus (isolated in 2009 from a human cutaneous lesion following transmission from domestic rats)*(24)*. Both strains were grown on Vero cells (Eagle’s Minimum Essential medium with 1% Penicillin-Streptomycin, 1% Glutamine and 10% foetal calf serum, 37°C with 5% CO_2_). Virus productions used for the sero-neutralization assay were optimized to obtain low and similar ratios of non-infectious particles to infectious particles. For vaccinia virus we used a culture supernatant harvested at D6 titrating to 1.04*10^9^ genome copies/mL and 1.67*10^6^ TCID50/mL (ratio genome copies/TCID50=624). For cowpox virus, we used an early supernatant collected at D2 titrating to 6.82*10^7^ genome copies/mL and 1.08*10^5^ TCID50/mL (ratio=613). Aliquots for individual experiments were prepared with 15mM HEPES buffer and frozen at -80°C.

The same seroneutralization protocol was used for both viruses. Dilutions of sera were performed in Eagle’s Minimum Essential medium with 1% Penicillin-Streptomycin in a 96-well format using an epMotion® 5075 (Eppendorf) working station. 50µL of dilution were mixed with the same volume of virus (50 TCID50/well) to produce final dilutions of 1:20, 1:40, 1:80 and 1:160. The plates were briefly centrifuged (300 rpm for 2 minutes) and incubated for 1 hour at 37°C. The serum/virus mixture was then plated into a cell culture 96-well plates containing confluent Vero cells and 100µL of culture medium (Eagle’s Minimum Essential medium with 1% Penicillin-Streptomycin, 1% Glutamine and 10% Fetal Calf Serum). Culture plates were incubated at 37°C under 5% CO_2_ for 4 days and included positive (serum of a donor multivaccinated with the Lister strain, supplied by the French National Reference Centre for Orthopoxviruses*(25))*.

Cytopathic effects (CPE) was observed at day 4 post infection. Pictures were obtained using the Cytation® (BioTek. Vermont, USA) or IncuCyte® (Sartorius. Gottingen, Germany) readers for subsequent analysis. Each sample was assigned a result: negative (default value: 1:10), or positive at 1:20, 1:40, 1:80 or 1:160 (corresponding to the highest dilution with no CPE)).

### Statistics

The distribution of serological titers and the proportion of positives and negatives were compared between decades of birth, regions or gender using the Mann-Whitney and Fisher tests, respectively.

The correlation between the anti-vaccinia and anti-cowpox neutralization titers was evaluated by a Spearman coefficient. Comparisons of distributions according to serological status were performed by Mann-Whitney tests for quantitative variables and Fisher tests for categorical variables. Seroprevalence was compared between each pair of regions by a Fisher test (without correction for test multiplicity). Factors associated with the serological titer were sought in univariate analysis and then after adjustment for year of birth by a parametric model based on the hypothesis of a lognormal distribution of titers and taking into account the interval censoring of serological titers*(26)*. For the French population, the covariates studied were the following: gender, marital status, type of occupation, level of education, number of people in the household, household income, general health status, travel outside Europe, type of housing, time spent outdoors, air conditioning, use of mosquito nets, presence of garden/terrace/balcony and swimming pool, rurality of the dwelling, proximity to shops, the presence of a pond or marsh nearby, contact with domestic or farm animals, exposure to mosquitoes and other biting insects and ticks, frequency of bites and protection used, type of water supply, contact with sewage, water consumption, contact with domestic or farm animals, hunting activity, type of meat consumed and cooking.

All tests were two-tailed at the 0.05 significance level.

## Results

### Antibodies neutralizing the vaccinia virus

Results for antibodies neutralizing the vaccinia virus in the different populations (Bolivia, Laos, Mali and France) are presented in Table 1. Overall seroprevalence was calculated for samples with a titer ≥40 (ThT40) or with a titer ≥20 (ThT20) (see discussion).

**Table 1:** Demographic characteristics and results of vaccinia virus neutralization in the 4 study populations.

|  | **BOLIVIA** | **LAOS** | **MALI** | **FRANCE** |
| --- | --- | --- | --- | --- |
| Number of participants | 601 | 657 | 255 | 4876 |
| Sex ratio m/f | 1.37 | 1.72 | 1.02 | 1.12 |
| Year of birth ± sd | 1989 ± 10 | 1985 ± 9 | 1980 ± 14 | 1967 ± 14 |
| Participants born before 1960 | 3 (0.5%) | 5 (0.8%) | 26 (10%) | 1560 (32%) |
| Participants born before 1980 | 161 (27%) | 171 (26%) | 112 (43%) | 3826 (78%) |
| Neutralizing titer <20 | 598 (99.50%) | 632 (96.19%) | 211 (82.74%) | 4137 (84.84%) |
| Neutralizing titer = 20 | 1 (0.17%) | 22 (3.35%) | 30 (11.76%) | 425 (8.72%) |
| Neutralizing titer = 40 | 1 (0.17%) | 3 (0.46%) | 12 (4.71%) | 217 (4.45%) |
| Neutralizing titer = 80 | 1 (0.17%) | 0 (0.00%) | 2 (0.78%) | 86 (1.76%) |
| Neutralizing titer = 160 | 0 (0.00%) | 0 (0.00%) | 0 (0.00%) | 11 (0.23%) |
| Overall seroprevalence ≥ 40 | **0.33%** | **0.46%** | **5.49%** | **6.44%** |
| Overall seroprevalence ≥ 20 | **0.50%** | **3.81%** | **17.25%** | **15.16%** |

### Cross immunity between vaccinia and cowpox virus

In the French population, 4448 samples from four regions had sufficient volumes to be tested for both vaccinia and cowpox viruses; 4391 (98.8%) had similar antibody titers for both viruses (same titer ± 1 dilution). Details are presented in Table 2. The correlation coefficient (Spearman) between the two serologies was estimated at 0.68, which supports the hypothesis of a high correlation between these titers.

**Table 2:** Cross-reactivity between neutralizing assays of vaccinia virus and cowpox virus.

|  |  | **Antibodies neutralizing cowpox virus** |  |  |  |  |
| --- | --- | --- | --- | --- | --- | --- |
|  |  | **10** | **20** | **40** | **80** | **160** |
| **Antibodies neutralizing vaccinia virus** | **10** | 3443<br>(77.44%) | 407<br>(9.15%) | 4<br>(0.09%) | 2<br>(0.04%) | 1<br>(0.02%) |
|  | **20** | 234<br>(5.26%) | 85<br>(1.91%) | 12<br>(0.27%) | 5<br>(0.11%) | 0<br>(0.00%) |
|  | **40** | 7<br>(0.16%) | 77<br>(1.73%) | 62<br>(1.39%) | 12<br>(0.27%) | 3<br>(0.07%) |
|  | **80** | 0<br>(0.00%) | 4<br>(0.09%) | 31<br>(0.70%) | 21<br>(0.47%) | 3<br>(0.07%) |
|  | **160** | 0<br>(0.00%) | 1<br>(0.02%) | 0<br>(0.00%) | 2<br>(0.04%) | 2<br>(0.04%) |

### Prevalence of antibodies neutralizing orthopoxviruses in French populations: epidemiological data by age, gender and region

Among the 4448 samples tested for both vaccinia and smallpox virus, sex ratio M/F was 1.17 and mean year of birth was 1966 ± 13 years). Given the difference observed between the seroprevalence values calculated using a threshold titer at 20 (ThT20) or 40 (ThT40) with the vaccinia microneutralization test (see above), we took advantage of the greater specificity offered by the double-testing procedure as follows: we calculated an “orthopoxviruses neutralization titer” (ONT) as the geometric mean of the vaccinia and smallpox neutralization titers and considered samples with an ONT titer ≥20 to be positive. Characteristics of the population studied according to age group and region are given in Table 3.

**Table 3:**
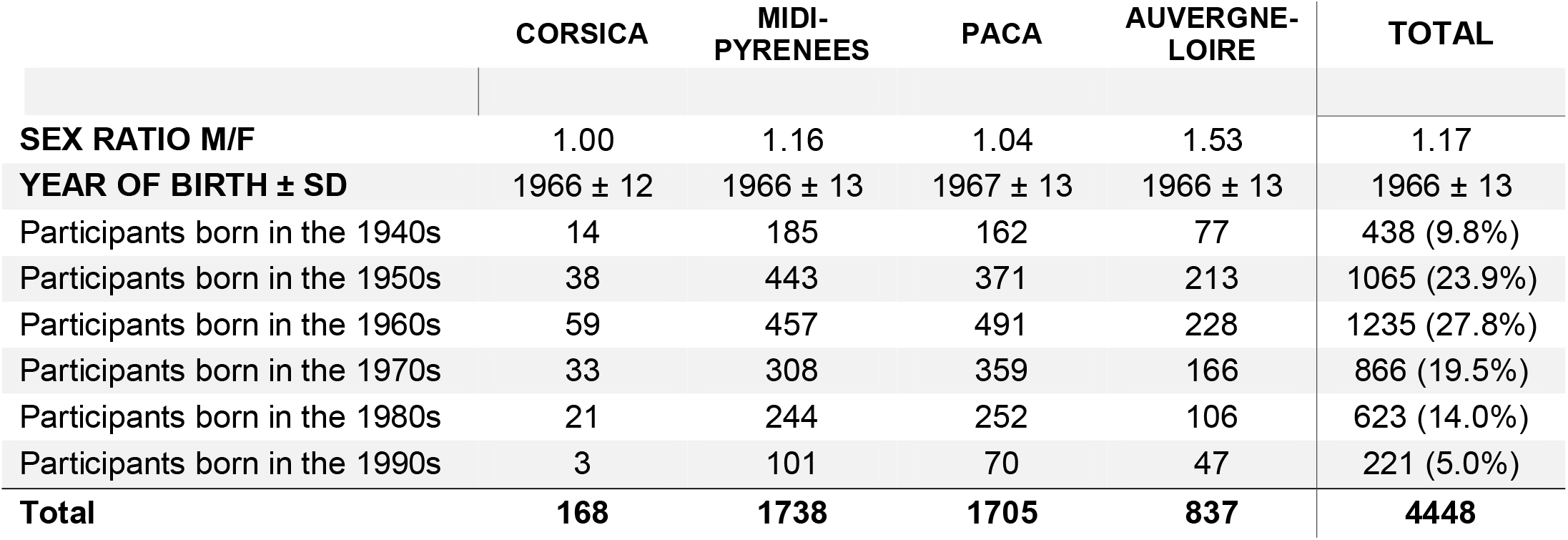
Demographic characteristics of the French population tested with both vaccinia and cowpox virus neutralization techniques. PACA: Provence Alpes Côte-d’Azur.

|  | CORSICA | MIDI-PYRENEES | PACA | AUVERGNE-LOIRE | TOTAL |
| --- | --- | --- | --- | --- | --- |
| **SEX RATIO M/F** | 1.00 | 1.16 | 1.04 | 1.53 | 1.17 |
| **YEAR OF BIRTH ± SD** | 1966 ± 12 | 1966 ± 13 | 1967 ± 13 | 1966 ± 13 | 1966 ± 13 |
| Participants born in the 1940s | 14 | 185 | 162 | 77 | 438 (9.8%) |
| Participants born in the 1950s | 38 | 443 | 371 | 213 | 1065 (23.9%) |
| Participants born in the 1960s | 59 | 457 | 491 | 228 | 1235 (27.8%) |
| Participants born in the 1970s | 33 | 308 | 359 | 166 | 866 (19.5%) |
| Participants born in the 1980s | 21 | 244 | 252 | 106 | 623 (14.0%) |
| Participants born in the 1990s | 3 | 101 | 70 | 47 | 221 (5.0%) |
| **Total** | **168** | **1738** | **1705** | **837** | **4448** |

The global seroprevalence was 8.18%. It was over 10% in individuals born before 1970, dropped to 5% in those born during the 1970’s, and to less than 1% after 1980. The geometric mean of ONT in the complete sample was 12.8. We observed limited differences according to age groups (Figure 1.a) but a clear overall increase of the proportion of positives (Figure 1.b) in relation with age. Antibody titers and percentages of positives per decade are given in figure 1.a and 1.b. The p-values of the tests comparing the neutralizing antibody titers and the proportion of positives between each decade of birth are given in supplemental Table 1.

**Figure 1:**
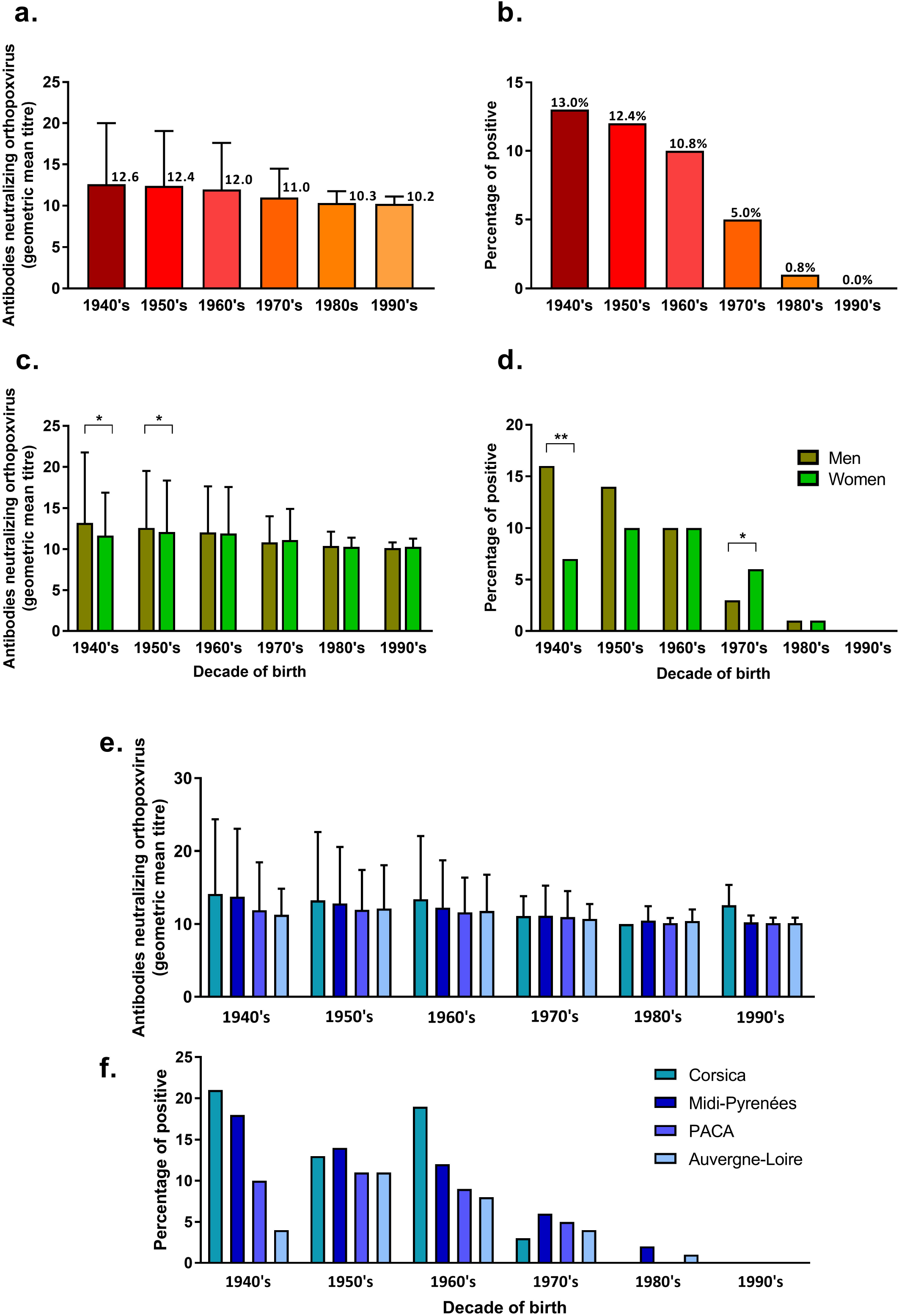
Results for the French population tested for both viruses (vaccinia and cowpox virus). Overall comparison of ONT (a.) and percentage of positives (ONT > 20) (b.) according to participants’ decade of birth (p-values are in Supplemental Table 1). Comparison for the same birth decade of the ONT (c.) and the percentage of positives (ONT > 20) (d.) by gender of the participants (Mann-Whitney and Fischer tests, respectively). Comparison for the same decade of ONT (e.) and percentage positive (ONT > 20) (f.) by living-region of participants (Mann-Whitney and Fischer tests, respectively).

The association between gender and antibodies neutralizing orthopoxviruses was different depending on the decade of birth. Until the 1950s, seroprevalence was higher among men, then balanced out in the 1960s and inverted in the 1970s when seroprevalence became higher among women. From the 1980s onwards, as there are hardly any more positive participants, no difference is observed (figure 1c. and 1d.).

The studied French sample geographically covers 4 regions: Auvergne-Loire (n=837 participants), Corsica (n=168), Midi-Pyrénées (n=1738) and PACA (n=1705). We compared the different regions for each decade. We observed a few differences between these 4 regions both in neutralizing antibodies titer and in percentages of positives (figures 1e. and 1f.). Observed seroprevalence was higher in Corsica and in the Midi-Pyrénées region than in the PACA region and especially the Auvergne-Loire region, which has the lowest seroprevalence of the four regions for each decade of birth.

Finally, during the collection of samples, French participants filled in a detailed questionnaire concerning lifestyle, eating habits, rurality index, contact with livestock or wild animals, etc. We estimated the association between each of these parameters and ONT. Sixty parameters were considered, and once the tests had been adjusted on age, none of the parameters were found to influence seroprevalence. In short, in our study the only parameter directly associated with the titer of orthopoxviruses neutralizing antibodies was age.

## Discussion

With the end of smallpox vaccination in the 1980s, it was expected that the prevalence of antibodies against orthopoxviruses would decline worldwide. Obviously, this seroprevalence would decrease in the youngest (and never vaccinated) age groups, but it could be assumed that the circulation of other orthopoxviruses would be responsible for the development of immunity by natural infection in some individuals. Similarly, while it is classically described that smallpox vaccination can produce long-lasting immunity, it could be assumed that the immunosenescence of older vaccinated populations would contribute to a decrease in seroprevalence in individuals vaccinated in childhood.

Consequently, the prevalence of anti-orthopoxvirus antibodies is difficult to anticipate precisely in a given population because the parameters that can modulate it are numerous (intensity of past vaccination campaigns, number of doses received, possible exposure to infection by orthopoxviruses, factors modulating immunosenescence etc.). Our study allows to provide concrete elements of seroprevalence and to compare several populations between them. Nevertheless, it has several limitations, such as the non-representativeness of the general population by the tested cohorts, obvious differences in the recruitment of participants in the different international populations, the absence of individual vaccine data, the weakness of the collected metadata, the limited number of elderly people tested and ultimately the absence of a strict internationally validated threshold value for neutralization tests.

Looking at the overall picture of the data collected in the current study, it is broadly in line with what was expected: seroprevalence is higher in countries that routinely vaccinated and in participants born before the cessation of smallpox vaccination. However, when one goes into more detail, a number of points deserve to be discussed.

The results in Laos and Bolivia are consistent with the previous WHO containment strategy used when vaccination of the general population was not feasible. The seroprevalence is remarkably low in Bolivia (<0.5%, whatever the threshold titer used) in both those born before and after 1980. This reflects both likely low vaccination coverage and lack of exposure to natural orthopoxvirus infection.

In Laos, the prevalence using a threshold titer of 40 (ThT40) is very low (<1%) but is close to 4% with a threshold titer of 20 (ThT20). The prevalence values for individuals born before 1980 are 1.75% (ThT40) and 7.02% (ThT20), compared with 0% and 2.67% for those born after 1980. Therefore, neutralizing antibodies are found predominantly (but not exclusively) in individuals born before 1980 and may be related to smallpox vaccination, although a low-level exposure to orthopoxvirus infection cannot be formally excluded.

In Mali, the ThT40 seroprevalence is ∼5% and rises to ∼17% for ThT20. The prevalence values for individuals born before 1980 are 10.71% (ThT40) and 27.68% (ThT20), compared with 1.38% and 8.97% for those born after 1980. Since health authorities confirmed that there was no smallpox vaccination campaign in the study area after 1980, the distribution of seroprevalence in age groups presented in Supplemental Figure 1 for ThT20 suggests that exposure to natural orthopoxvirus infection accounts for part of the observed immunity. This is consistent with the fact that the samples were collected in villages located in a forested area (southern Mali), and that circulation of monkeypox in humans, monkeys, and rodents has been reported in central and western Africa for several decades, primarily at the edge of forests*(3)*. However, it is clear that vaccination may also explain the specifically high prevalence in the oldest age groups in a proportion that we cannot determine.

In France, we had the opportunity to perform a double orthopox testing (vaccinia virus and cowpox virus) in a large part of the population studied to improve the specificity of the neutralization procedure. Results for both viruses were very similar (98% of participants had concordant titers in both tests), confirming extensive cross-immunity within the orthopoxvirus genus, which is well documented for smallpox, vaccinia and monkeypox viruses*(27)*. We found a global seroprevalence of 8.18%, which is mainly related to age (and thus to smallpox vaccination coverage), with a sharp decline from the 1970s. We observed gender differences, especially in the elderly, where it is higher in men (see Figure 1c. and 1d.). We assume that the medical rigour resulting from military service for men in France may have played a role in this age group.

In our study, we observed that some participants born in the early 1940s still have high neutralization titers for orthopoxviruses. This is consistent with previous reports that smallpox vaccination generates long-term splenic memory lymphocytes that can lead to the production of anti-vaccine antibodies more than 80 years after vaccination*(28–37)*. However, our results also show a low seroprevalence among participants born before 1960 (∼12.5%) compared to the vaccination coverage estimated at 90% by Santé Publique France among people born before 1966*(20)*. These data suggest that individuals show, in proportions that are difficult to define but higher than those suggested in the literature*(34,38)*, a decrease in antibodies. To what extent previously vaccinated individuals in whom no neutralizing antibody can be detected retain any form of functional immunity against orthopoxviruses is unknown.

Our results also show that compliance with the smallpox vaccine or booster shots in France had declined well before the end of compulsory vaccination with possible territorial disparities. Smallpox had disappeared from Europe since the First World War and the epidemics generated by imported cases in the 1950s*(39,40)* consistently suggest that vaccination coverage had already begun to decline, driven by the adverse effects of the vaccine.

Interestingly, ThT20 prevalence values in individuals born after 1980 are very low (1.74% in Midi-Pyrenées, 0.65% in Auvergne-Loire and 0% in PACA), suggesting the absence of natural orthopoxvirus infections. Further investigations and a larger number of individuals tested are needed to decide on the case of Corsica where we identified a proportion of individuals born after 1980 with antibodies to vaccinia virus. No environmental factors were associated with seroprevalence in the French population despite a large database on living conditions.

In conclusion, our study suggests that, overall, the different populations that we have tested in Europe, Africa, Asia and South America are massively susceptible to orthopoxvirus infections. Even in Africa, where there is significant evidence of natural circulation of orthopoxviruses, population immunity is modest. Levels of protection are the lowest in individuals born after 1980, due to the end of smallpox vaccination four decades ago. In practical terms, this means that there is no population immunity to provide a barrier to the spread of orthopoxviruses, and fully confirms that the cessation of smallpox vaccination may facilitate the emergence of orthopoxvirus infections, as currently observed in the spread of monkeypox virus in Europe.

## Data Availability

All data produced in the present study are available upon reasonable request to the authors

## Biographical sketch

Dr. Luciani is a pharmacist medical biologist working at Assistance Publique Hôpitaux de Marseille and at the Unité des Virus Emergents, Aix Marseille Université, Marseille, France. Her primary research interests include poxviruses and diagnostics of emerging viral infections.

## Acknowledgments

This work was supported by the European Union’s Horizon 2020 Research and Innovation Programme under grant agreement 653316 (European Virus Archive goes global project: http://www.european-virus-archive.com/). We are very grateful to the donors, and the staff of the National Blood Transfusion Centre of the Lao Red Cross. We thank the staff of Microbiology Laboratory, Mahosot Hospital, Dr Manivanh Vongsouvath (director), and Dr Koukeo Phommasone (deputy director). We thank the staff of LOMWRU (Lao-Oxford-Mahosot Hospital-Wellcome Trust-Research Unit), and Prof. Paul N Newton (former director).

## References

1. Beer EM, Rao VB. A systematic review of the epidemiology of human monkeypox outbreaks and implications for outbreak strategy. PLoS Negl Trop Dis [Internet]. 2019 Oct 16 [cited 2020 Apr 15];13(10). Available from: https://www.ncbi.nlm.nih.gov/pmc/articles/PMC6816577/

2. Doshi RH, Guagliardo SAJ, Doty JB, Babeaux AD, Matheny A, Burgado J, et al. Epidemiologic and Ecologic Investigations of Monkeypox, Likouala Department, Republic of the Congo, 2017. Emerg Infect Dis. 2019 Feb;25(2):281–9.

3. Durski KN, McCollum AM, Nakazawa Y, Petersen BW, Reynolds MG, Briand S, et al. Emergence of Monkeypox — West and Central Africa, 1970–2017. MMWR Morb Mortal Wkly Rep. 2018 Mar 16;67(10):306–10.

4. Centers for Disease Control and Prevention (CDC). Update: multistate outbreak of monkeypox--Illinois, Indiana, Kansas, Missouri, Ohio, and Wisconsin, 2003. MMWR Morb Mortal Wkly Rep. 2003 Jun 27;52(25):589–90.

5. Vaughan A, Aarons E, Astbury J, Balasegaram S, Beadsworth M, Beck CR, et al. Two cases of monkeypox imported to the United Kingdom, September 2018. Eurosurveillance [Internet]. 2018 Sep 20 [cited 2019 Feb 8];23(38). Available from: https://www.eurosurveillance.org/content/10.2807/1560-7917.ES.2018.23.38.1800509

6. Venkatesan G, Balamurugan V, Prabhu M, Yogisharadhya R, Bora DP, Gandhale PN, et al. Emerging and reLemerging zoonotic buffalopox infection: a severe outbreak in Kolhapur (Maharashtra), India. Vet Ital. 2010;46:10.

7. Kolhapure RM, Deolankar RP, Tupe CD, Raut CG, Basu A, Dama BM, et al. Investigation of buffalopox outbreaks in Maharashtra State during 1992-1996. Indian J Med Res. 1997 Nov;106:441–6.

8. Prabhu M. Camelpox and Buffalopox: Two Emerging and Re-emerging Orthopox Viral Diseases of India. Adv Anim Vet Sci. 2015;3(10):527–41.

9. Balamurugan V, Venkatesan G, Bhanuprakash V, Singh RK. Camelpox, an emerging orthopox viral disease. Indian J Virol. 2013 Dec 1;24(3):295–305.

10. Bera BC, Shanmugasundaram K, Barua S, Venkatesan G, Virmani N, Riyesh T, et al. Zoonotic cases of camelpox infection in India. Vet Microbiol. 2011 Aug;152(1–2):29–38.

11. Duraffour S, Mertens B, Meyer H, van den Oord JJ, Mitera T, Matthys P, et al. Emergence of Cowpox: Study of the Virulence of Clinical Strains and Evaluation of Antivirals. Tavis JE, editor. PLoS ONE. 2013 Feb 15;8(2):e55808.

12. Kurth A, Wibbelt G, Gerber HP, Petschaelis A, Pauli G, Nitsche A. Rat-to-Elephant-to-Human Transmission of Cowpox Virus. Emerg Infect Dis. 2008 Apr;14(4):670–1.

13. Wolfs TFW, Wagenaar JA, Niesters HGM, Osterhaus ADME. Rat-to-Human Transmission of Cowpox Infection. Emerg Infect Dis. 2002 Dec;8(12):1495–6.

14. Keesing F, Belden LK, Daszak P, Dobson A, Harvell CD, Holt RD, et al. Impacts of biodiversity on the emergence and transmission of infectious diseases. Nature. 2010 Dec;468(7324):647–52.

15. Keesing F, Ostfeld RS. Impacts of biodiversity and biodiversity loss on zoonotic diseases. Proc Natl Acad Sci [Internet]. 2021 Apr 27 [cited 2021 Sep 16];118(17). Available from: https://www.pnas.org/content/118/17/e2023540118

16. Schmeller DS, Courchamp F, Killeen G. Biodiversity loss, emerging pathogens and human health risks. Biodivers Conserv. 2020 Oct 1;29(11):3095–102.

17. Berche P. The threat of smallpox and bioterrorism. Trends Microbiol. 2001 Jan 1;9(1):15–8.

18. Cohen J. BIOTERRORISM: Smallpox Vaccinations: How Much Protection Remains? Science. 2001 Nov 2;294(5544):985–985.

19. Lane HC, Montagne JL, Fauci AS. Bioterrorism: A clear and present danger. Nat Med. 2001 Dec 1;7(12):nm1201-1271–1271.

20. Lévy-Bruhl D, Guérin N. Utilisation du virus de la variole comme arme biologiqueL: place de la vaccination en France. Eurosurveillance. 2001 Nov 1;6(11):171–8.

21. Fenner F, Henderson DA, Arita I, Jezek Z, Ladnyi ID, Organization WH. Smallpox and its eradication. Geneva: GenevaL: World Health Organization; 1988.

22. Poinsignon A, Boulanger D, Binetruy F, Elguero E, Darriet F, Gallian P, et al. Risk factors of exposure to Aedes albopictus bites in mainland France using an immunological biomarker. Epidemiol Infect [Internet]. 2019 ed [cited 2021 Sep 13];147. Available from: https://www.cambridge.org/core/journals/epidemiology-and-infection/article/risk-factors-of-exposure-to-aedes-albopictus-bites-in-mainland-france-using-an-immunological-biomarker/45001FFB8F03EA609059592B005211DA

23. Saba Villarroel PM, Nurtop E, Pastorino B, Roca Y, Drexler JF, Gallian P, et al. Zika virus epidemiology in Bolivia: A seroprevalence study in volunteer blood donors. PLoS Negl Trop Dis. 2018 Mar 7;12(3):e0006239.

24. Ninove L, Domart Y, Vervel C, Voinot C, Salez N, Raoult D, et al. Cowpox Virus Transmission from Pet Rats to Humans, France. Emerg Infect Dis. 2009 May;15(5):781–4.

25. Leparc-Goffart I, Poirier B, Garin D, Tissier MH, Fuchs F, Crance JM. Standardization of a neutralizing anti-vaccinia antibodies titration method: an essential step for titration of vaccinia immunoglobulins and smallpox vaccines evaluation. J Clin Virol. 2005 Jan;32(1):47–52.

26. Lapidus N, Lamballerie X de Salez N, Setbon M, Delabre RM, Ferrari P, et al. Factors Associated with Post-Seasonal Serological Titer and Risk Factors for Infection with the Pandemic A/H1N1 Virus in the French General Population. PLOS ONE. 2013 Apr 16;8(4):e60127.

27. Edghill-Smith Y, Golding H, Manischewitz J, King LR, Scott D, Bray M, et al. Smallpox vaccine–induced antibodies are necessary and sufficient for protection against monkeypox virus. Nat Med. 2005 Jul;11(7):740–7.

28. El-Ad B, Roth Y, Winder A, Tochner Z, Lublin-Tennenbaum T, Katz E, et al. The Persistence of Neutralizing Antibodies after Revaccination against Smallpox. J Infect Dis. 1990 Mar 1;161(3):446–8.

29. Demkowicz WE, Littaua RA, Wang J, Ennis FA. Human cytotoxic T-cell memory: long-lived responses to vaccinia virus. J Virol. 1996 Apr;70(4):2627–31.

30. Crotty S, Felgner P, Davies H, Glidewell J, Villarreal L, Ahmed R. Cutting Edge: Long-Term B Cell Memory in Humans after Smallpox Vaccination. J Immunol. 2003 Nov 15;171(10):4969–73.

31. Pütz MM, Alberini I, Midgley CM, Manini I, Montomoli E, Smith GLY 2005. Prevalence of antibodies to Vaccinia virus after smallpox vaccination in Italy. J Gen Virol. 86(11):2955–60.

32. Hammarlund E, Lewis MW, Hansen SG, Strelow LI, Nelson JA, Sexton GJ, et al. Duration of antiviral immunity after smallpox vaccination. Nat Med. 2003 Sep;9(9):1131–7.

33. Combadiere B, Boissonnas A, Carcelain G, Lefranc E, Samri A, Bricaire F, et al. Distinct Time Effects of Vaccination on Long-Term Proliferative and IFN-γ–producing T Cell Memory to Smallpox in Humans. J Exp Med. 2004 Jun 7;199(11):1585–93.

34. Taub DD, Ershler WB, Janowski M, Artz A, Key ML, McKelvey J, et al. Immunity from Smallpox Vaccine Persists for Decades: A Longitudinal Study. Am J Med. 2008 Dec 1;121(12):1058–64.

35. Sarkar JK, Mitra AC, Mukherjee MK. The minimum protective level of antibodies in smallpox. Bull World Health Organ. 1975;52(3):307–11.

36. Amanna IJ, Carlson NE, Slifka MK. Duration of Humoral Immunity to Common Viral and Vaccine Antigens. N Engl J Med. 2007 Nov 8;357(19):1903–15.

37. Mamani-Matsuda M, Cosma A, Weller S, Faili A, Staib C, Garçon L, et al. The human spleen is a major reservoir for long-lived vaccinia virus–specific memory B cells. Blood. 2008 May 1;111(9):4653–9.

38. Anis E, Leventhal A, Slater PE, Shinar E, Yahalom V, Smetana Z, et al. Smallpox revaccination of 21000 first responders in Israel: lessons learned. Int J Infect Dis. 2009 May 1;13(3):403–9.

39. Biraben JN. La diffusion de la vaccination en France au XIXe siècle. Ann Bretagne Pays Ouest. 1979;86(2):265–76.

40. Le Boukdelles B. The Smallpox Epidemic of 1955 in France. Bull Académie Natl Médecine. 1955;139(25/26):417–20.

