## Supplemental data for "Susceptibility to monkeypox virus infection: seroprevalence of orthopoxvirus in 4 population samples; France, Bolivia, Laos and Mali"

| **Antibodies neutralising orthopoxviruses** | | | | | |  | **Proportion of positive participants** | | | | | |
| --- | --- | --- | --- | --- | --- | --- | --- | --- | --- | --- | --- | --- |
|  | **1940's** | **1950's** | **1960's** | **1970's** | **1980's** |  |  | **1940's** | **1950's** | **1960's** | **1970's** | **1980's** |
| **1950's** | 0,856 |  |  |  |  |  | **1950's** | 0,733 |  |  |  |  |
| **1960's** | 0,082 | 0,039 |  |  |  |  | **1960's** | 0,131 | 0,113 |  |  |  |
| **1970's** | <0,0001 | <0,0001 | <0,0001 |  |  |  | **1970's** | <0,0001 | <0,0001 | <0,0001 |  |  |
| **1980's** | <0,0001 | <0,0001 | <0,0001 | <0,0001 |  |  | **1980's** | <0,0001 | <0,0001 | <0,0001 | <0,0001 |  |
| **1990's** | <0,0001 | <0,0001 | <0,0001 | 0,0002 | 0,535 |  | **1990's** | <0,0001 | <0,0001 | <0,0001 | <0,0001 | 0,334 |

**Supplemental Table 1:** p-values associated to Figure 1a. and 1b. Comparison of ONT and proportion of positive between decades of birth using a Mann-Whitney test.

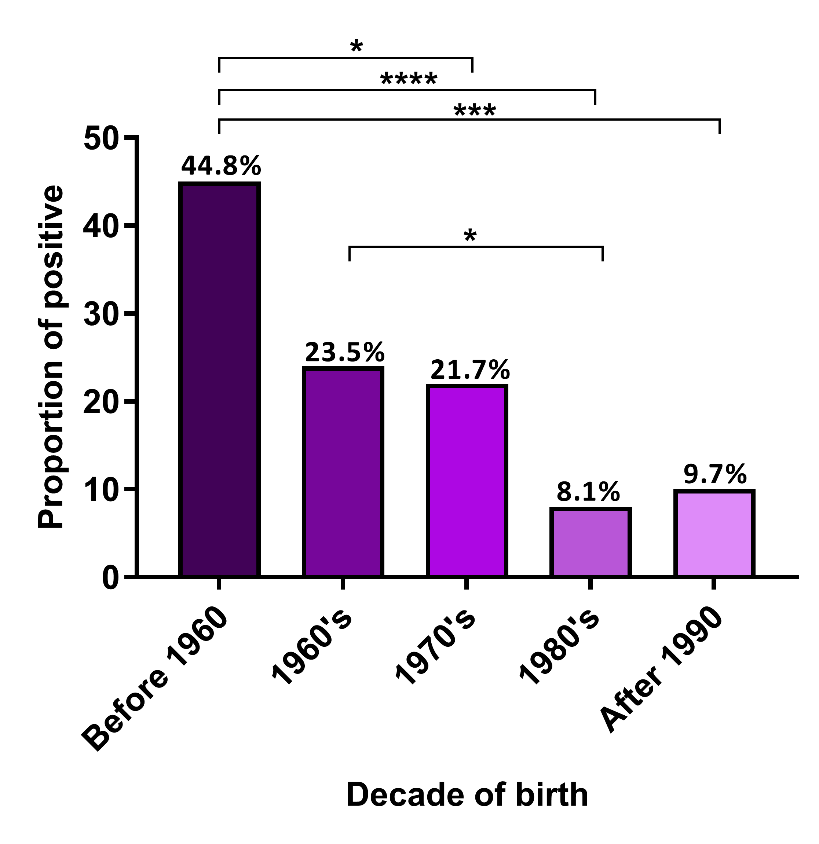

**Supplemental Figure 1**: seroprevalence in Mali using a threshold titre at 20 (ThT20). The numbers in each group are as follows: 29 for participants born before 1960, 34 for those born in the 1960s, 46 for those born in the 1970s, 74 for those born in the 1980s and 72 for those born after 1990.
